# SHELTER IN PLACE ORDER CONTAINED COVID-19 GROWTH RATE IN GREECE

**DOI:** 10.1101/2020.06.08.20125666

**Authors:** T. Giannouchos, A. Giannouchos, I. Christodoulou, E. Steletou, K. Souliotis

**Affiliations:** Pharmacotherapy Outcomes Research Center, College of Pharmacy, University of Utah, Salt Lake City, USA; Laboratory of Health Economics, University of Piraeus, Greece; St. Vinzenz Hospital, Dusseldorf, Germany; Department of Oncology, Johns Hopkins School of Medicine, Baltimore, USA; Department of Pediatrics, Regional General University Hospital of Patras, Greece; Department of Social and Education Policy, University of Peloponnese, Corinth, Greece

## Abstract

**Background:** The Greek authorities implemented the strong social distancing measures within the first few weeks after the first confirmed case of the virus to curtail the COVID-19 growth rate.

**Objectives:** To estimate the effect of the two-stage strong social distancing measures, the closure of all non-essential shopping centers and businesses on March 16 and the shelter in place orders (SIPOs) on March 23 on the COVID-19 growth rate in Greece

**Methods:** We obtained data on COVID-19 cases in Greece from February 26^th^ through May 4^th^ from publicly available sources. An interrupted time-series regression analysis was used to estimate the effect of the measures on the exponential growth of confirmed COVID-19 cases, controlling for the number of daily testing, and weekly fixed-effects.

**Results:** The growth rate of the COVID-19 cases in the pre-policies implementation period was positive as expected (p=0.003). Based on the estimates of the interrupted time-series, our results indicate that the SIPO on March 23 significantly slowed the growth rate of COVID-19 in Greece (p=0.04). However, we did not find evidence on the effectiveness of standalone and partial measures such as the non-essential business closures implemented on March 16 on the COVID-19 spread reduction.

**Discussion:** The combined social distancing measures implemented by the Greek authorities within the first few weeks after the first confirmed case of the virus reduced the COVID-19 growth rate. These findings provide evidence and highlight the effectiveness of these measures to flatten the curve and to slow the spread of the virus.

## Introduction

The rapid transmission of the coronavirus disease 2019 (COVID-19) from person-to-person resulted in the uptake of aggressive medical and public health responses as provisions to curtail the spread of the virus across multiple countries.^1,2^ The most notable of these policies included imposing shelter in place orders (SIPOs), closing public schools and non-essential businesses, avoiding the use of public transport and banning large social gatherings. Early evidence suggests that the adoption, implementation of and compliance with social distancing policies reduced traveling and movement of people and were thus effective strategies to reduce the COVID-19 exponential growth rate.^3–5^

Greece has a constrained healthcare system capacity and was particularly vulnerable to the COVID-19 pandemic due to its limited and scarce healthcare resources such as the low number of Intensive Care Unit (ICU) beds.^6^ Despite, Greece totals only 2834 confirmed cases and 163 deaths as of May 17^th^ overall, and is a successful outlier to the current global trends.^7^ The first laboratory-confirmed case was reported on February 26^th^ and the first death on March 12^th^.^8^ The Greek government immediately imposed policies to address the spread of the virus. In particular, on March 16^th^ all non-essential shopping centers and businesses closed and on March 23^rd^ stricter measures including SIPOs and penalties for violators were imposed, effective until May 4^th^.

Beyond these measures, the Greek government simultaneously initiated media campaigns to promote protective behaviors, such as hand washing and wearing masks, and daily updated the public on COVID-19 related outcomes and risks. Hence, a critical issue is whether the early government policies and restrictions tailored at mitigating the spread of the virus mainly through social distancing were effective to reduce its spread relative to other information-related provisions.

The objective of this study was to estimate the impact of the two-stage social distancing measures implemented by the Greek government from February 26^th^ through May 4^th^ on the growth rate of the confirmed COVID-19 cases. This study contributes to the current literature on social distancing measures’ intended outcomes and provides evidence on the effectiveness and the benefits of such measures on flattening the curve and slowing the spread of COVID-19.

## Data and Methods

We obtained data on COVID-19 cases in Greece from February 26^th^ through May 4^th^ from publicly available sources.^8^ These days were selected, as they indicate the period between the first confirmed COVID-19 case (26 February) and the lift of SIPOs (4 May). The outcome of interest was the daily growth rate in confirmed COVID-19 cases at the country level.

We calculated growth rates as the daily difference between the confirmed cases, which resulted in 69 observations-days. Since most epidemiological models use the cumulative natural log, we used this form to calculate the daily growth rate. Consistent with previous work, we added one to case counts to avoid dropping days with zero COVID-19 cases.^3^

We used interrupted time-series regression analysis to isolate and to estimate the effect of the measures on the exponential growth of confirmed COVID-19 cases. The interrupted time series is a strong quasi-experimental alternative commonly used for evaluating healthcare outcomes in the absence of comparison groups when the specific point in time at which the policy was implemented is known.^9,10^ If the policies had an impact, then the causal hypothesis is that the COVID-19 growth rate will exhibit a significantly different slope or level after the policies went into effect.

Since the number of confirmed cases might be conditional and correlated to the number of COVID-19 tests, we included daily performed tests as a control variable. In addition, because additional measures targeting specific geographical areas and populations were implemented weekly, we included week fixed effects to adjust for these provisions. To ensure that our model accounts for autocorrelation, which is common when using time-series data, we performed a test proposed by Cumby and Huizinga and adjusted accordingly.^11^ Finally, due to the incubation and pre-symptomatic period and the time lag between symptoms and COVID-19 confirmation, we conducted multiple robustness checks by setting the policy implementation periods two and four days later. All analyses were conducted using Stata version 16.1 (StataCorp, College Station, TX).

### Limitations

Multiple confounders might bias our findings particularly if other unobserved interventions occurred simultaneously that were not adjusted for, such as nonessential transport bans, night curfew, and government information campaigns. However, most of these provisions were implemented within a few days from one another and are highly correlated. Thus, it is not possible to identify the effect of each. Nonetheless, we control for week fixed effects and pre-policy implementation trends, which we believe sufficiently address these threats to causality.^3^ Also, due to the asymptomatic and pre-symptomatic nature of the virus coupled with the relative limited tests (around 1% of the total population), current COVID-19 cases might understate the true infection rate and the prevalence of the disease in the population.^12^

## Results

Overall, 2632 laboratory COVID-19 cases were confirmed from February 26 through May 4 in Greece. The overall distribution of cumulative COVID-19 cases is presented in Figure 1.

**Figure 1:**
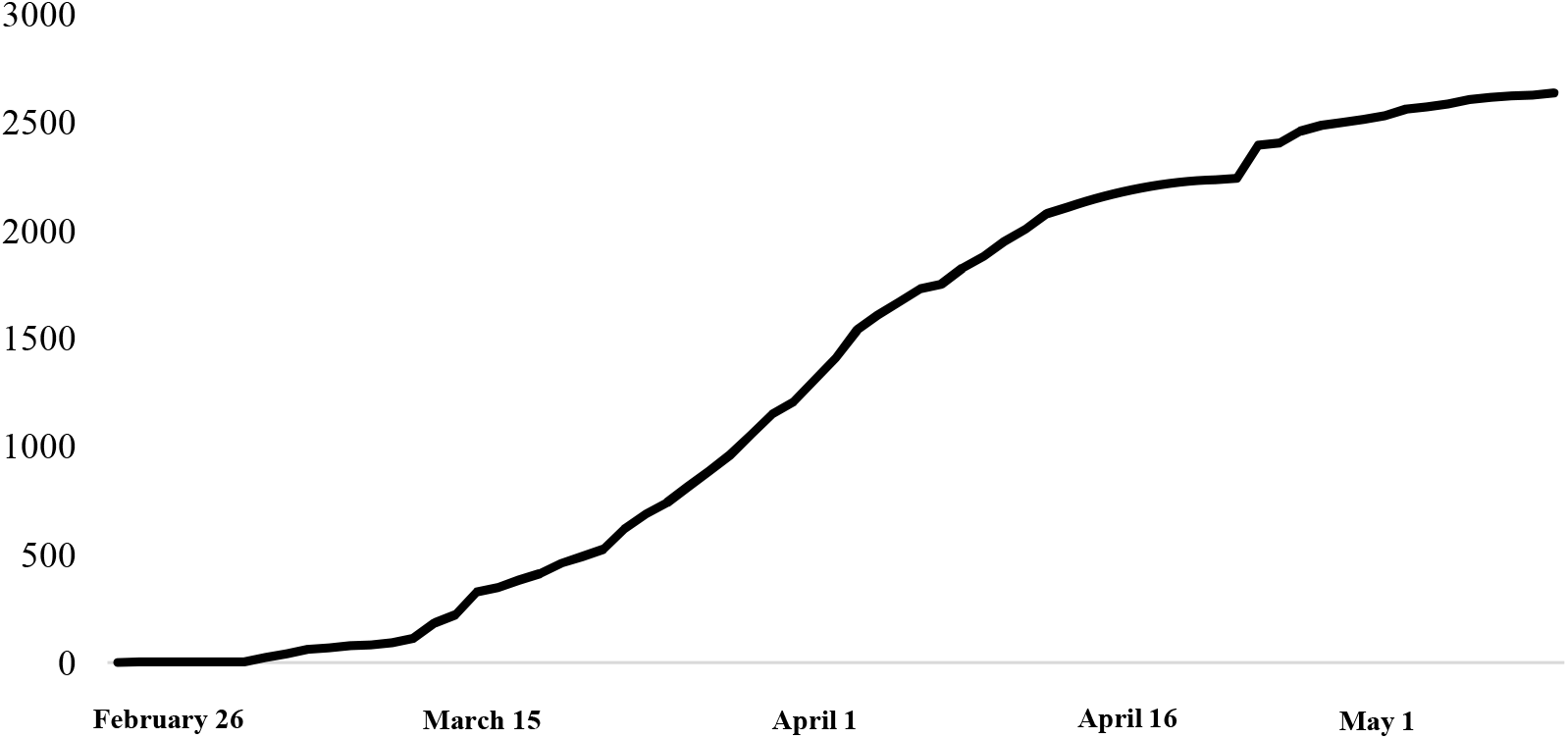
Cumulative COVID-19 cases in Greece until May 4^th^

Estimates of the interrupted time series regression on the social distancing policies’ impact on COVID-19 daily growth rate are presented in Table 1.

**Table 1:** Interrupted time series regression analyses estimates of social distancing policies on COVID-19 growth rates

|  | **Coefficient (95% CI)** | **p-value** |
| --- | --- | --- |
| **Pre-policies growth rate** | 0.15 ( 0.05 - 0.24) | 0.003 |
| **Non-essential shopping centers closure (March 16)** | -0.90 (-1.54 - -0.26) | 0.006 |
| **Growth rate after March 16** | -0.08 (-0.25 - 0.08) | 0.317 |
| **Shelter in place order (March 23)** | 0.53 (-0.33 - 1.40) | 0.225 |
| **Growth rate after March 23** | -0.17 (-0.33 - -0.07) | 0.040 |

The daily growth rate was positive and statistically significant in the pre-policies implementation period, as expected. The measure related to the closure of non-essential shopping centers implemented on March 16 yielded an insignificant change in the COVID-19 confirmed cases over the week between March 16 and March 23, which preceded the SIPO measures (p=0.317). However, we found a statistically significant reduction in the growth rate of COVID-19 cases in the time period after March 23 when the SIPO measures went into effect (p<0.05) (Figure 2). Our sensitivity analyses using two- and four-days time-lags yielded similar findings, thus corroborating the robustness of our findings.

**Figure 2:**
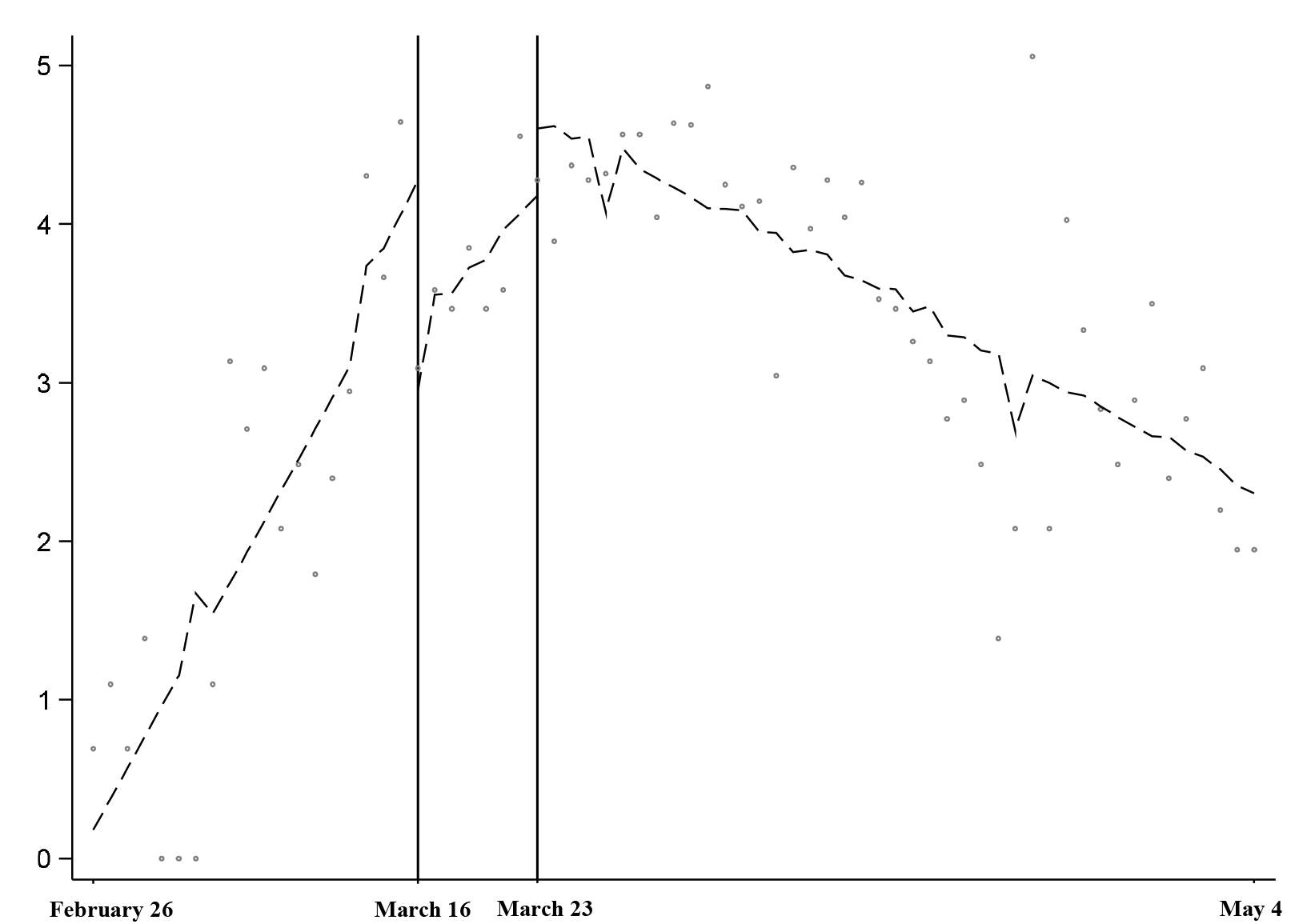
Growth rates before and after the social distancing policies’ implementation based on the interrupted time series estimates

## Discussion

Our results indicate that the shelter-in-place order on March 23 slowed the growth rate of COVID-19 in Greece. Had the Greek government not acted rapidly to contain the COVID-19 spread, the number of COVID-10 cases, and the related hospitalizations and deaths would have been exponentially higher. However, we did not find evidence on the effectiveness of standalone and partial measures such as the non-essential business closures implemented on March 16 on the COVID-19 spread reduction.

The findings regarding the effectiveness of strict social distancing shelter-in-place measures corroborate and give context to the existing evidence on Greece’s success in containing the spread of the virus and suggest that the rapid governmental response had the desired outcome on flattening the outbreak’s growth curve. Our results are also supported by a recent study, which estimated that 4360 deaths were averted in Greece by April 27^th^, as a result of early governmental interventions.^13^ Our estimates are also in-line with findings in the United States, where the implementation of SIPOs reduced the daily growth of the COVID-19 spread by almost 35 times by April 27.^3^

The lack of evidence on the impact of the ban on non-essential business operations suggests that this policy did not reduce social interactions, but rather displaced and replaced them with informal events and gatherings. For example, community mobility report data indicate a 24% to 53% increase in visits to public spaces through May 9 in Greece, compared to a baseline period from January 3 to February 6.^14^ Similar insignificant effects of such policies were also found in the United States.^3^

The European Union Council underlined the need to not only limit the spread of the virus but also to expand the capacity and address inefficiencies of the healthcare system, to promote research and to manage the socioeconomic impact of the virus.^15^ To this, we also highlight the importance of transparent communication with the public and income assurances, which are critical to maintain the public’s trust in public health authorities and political leaders and to reduce hardship.^16^ Providing financial assistance and a steady income flow during the quarantine was a crucial determinant of compliance to social distancing measures in Israel, which indicates additional considerations and trade-offs that official authorities should take into account.^17^

Our results evaluate the impact of shelter in place orders and other social distancing measures on the spread of COVID-19 in Greece. Our contribution aims to provide credible evidence about the effectiveness of strong social distancing measures that could guide policymakers in the face of adverse outcomes due to the de-escalation of these measures. Future research may examine the effect of de-escalation of such measures from May 4 onwards on COVID-19 containment.

## Data Availability

DATA ARE PUBLICLY AVAILABLE ONLINE AND SOURCE IS CITED IN THE MANUSCRIPT

